# Compositional structural brain signatures capture Alzheimer’s genetic risk on brain structure along the disease *continuum*

**DOI:** 10.1101/2024.05.08.24307046

**Authors:** Patricia Genius, M.Luz Calle, Blanca Rodríguez-Fernández, Carolina Minguillon, Raffaele Cacciaglia, Diego Garrido-Martin, Manel Esteller, Arcadi Navarro, Juan Domingo Gispert, Natalia Vilor-Tejedor, Alzheimer’s Disease Neuroimaging Initiative, ALFA study

## Abstract

**INTRODUCTION:** Traditional brain imaging genetics studies have primarily focused on how genetic factors influence the volume of specific brain regions, often neglecting the overall complexity of brain architecture and its genetic underpinnings.

**METHODS:** This study analyzed data from participants across the Alzheimer’s disease (AD) *continuum* from the ALFA and ADNI studies. We exploited compositional data analysis to examine relative brain volumetric variations that (i) differentiate cognitively unimpaired (CU) individuals, defined as amyloid-negative (A-) based on CSF profiling, from those at different AD stages, and (ii) associated with increased genetic susceptibility to AD, assessed using polygenic risk scores.

**RESULTS:** Distinct brain signatures differentiated CU A-individuals from amyloid-positive MCI and AD. Moreover, disease stage-specific signatures were associated with higher genetic risk of AD.

**DISCUSSION:** The findings underscore the complex interplay between genetics and disease stages in shaping brain structure, which could inform targeted preventive strategies and interventions in preclinical AD.

## 1. Background

Alzheimer’s disease (AD) is a complex multifactorial and heterogeneous neurodegenerative disorder, characterized by progressive changes in brain structure, among other pathophysiological hallmarks^1^. Structural magnetic resonance imaging (MRI) serves as a powerful tool for quantifying cerebral atrophy in the AD *continuum*. Common structural brain changes in AD display a limbic-to-frontal sequence^2^ throughout the disease *continuum*. Notably, brain atrophy and hippocampal reduction are closely linked with cognitive decline ^3–4–5–6^. Recent studies have shown the high heritability of these structural changes ^7–8^, which increases the interest in their role as intermediate phenotypes. Genetics plays a pivotal role in AD, with the *APOLIPOPROTEIN E* gene allele *ε4* (*APOE-ε4*) significantly increasing disease risk^9^. Nonetheless, beyond *APOE-ε4*, genome-wide association studies (GWAS) have identified numerous genetic variants associated with AD ^10–11–12^. In a recent published study ^13^ we showed that the genetic risk of AD, estimated through polygenic risk scoring, was higher in clinical groups than in cognitively unimpaired (CU) amyloid negative individuals, but not amyloid positive. These findings highlight a similar genetic burden between CU individuals displaying AD-related pathology and individuals at clinical stages of the disease. Therefore, these results reinforce the need to carefully examine how genetics impact on AD structural endophenotypes to examine brain cortical and subcortical changes not only based on the clinical diagnosis but also on the genetic load for AD at early stages of the disease. By combining imaging and genetic markers, there is potential for a more comprehensive exploration and understanding of the individuals’ profiles during the preclinical stages of the disease. In this line, brain imaging genetics (IG) studies aim to integrate neuroimaging (NI) and genetic data to uncover new genetic variants associated with AD-related brain features ^14–15^. Among the different approaches in IG, univariate analyses that look for the association between individual brain volumetric structures and AD genetic risk factors ^16–18^, do not consider the compositional origin of the brain. The brain can be segmented in regions, and the volumes of the segmented regions can be expressed as a fraction relative to the total intracranial volume. Similarly, at any level of segmentation, a specific brain area and its segmented regions can be defined as a composition and its components, respectively. Thus, given the interconnected and compositional structure of the brain, it is crucial to analyze brain features in a multivariate way to fully understand the complex interactions and influences across different regions^19^. Recent studies have used compositional approaches in the field of brain imaging due to the compositional structure of the data obtained from structural MRI and anatomical brain segmentation ^20^. Compositional data analysis (CoDA) methods offer a sophisticated approach to evaluating the brain’s complex structure, by viewing it as a cohesive entity. In CoDA, instead of exploring each component separately, the analysis focuses on the relative variation between them ^21–22^. This approach moves beyond the limitations of traditional analyses that either focus on single volumetric structures or employ multivariate techniques that fail to capture the compositional origin of structural brain features. This aspect is particularly crucial in AD, where changes in one region can have cascading effects on others, reflecting the multifaceted nature of the disease^23^. This study aimed to identify relative brain volumetric variations in cortical and subcortical areas that (i) distinguished between CU individuals and others at different stages of the disease and (ii) were associated with higher genetic susceptibility to AD within groups along the *continuum*. Compositional methods were employed to explore these relative brain volumetric variations, summarized in structural brain signatures. Results showed that structural brain signatures were accurately distinguishing between CU individuals and others at different stages of the disease. Moreover, disease stage-specific signatures were associated with higher genetic risk of AD. These findings highlight the modifying effect of disease stage and the heterogeneous profiles within the *continuum* on how AD genetic burden impacts brain structure, demonstrating the complex interplay between genetics and disease stage in shaping structural brain volumetry.

## 2. Methods

### 2.1. Study Population

This study included 338 CU middle-aged (45-65 years old) participants from the ALFA+ cohort, a nested cohort of the ALzheimer’s and FAmilies study^24^, and 330 CI individuals from the Alzheimer’s Disease Neuroimaging Initiative (ADNI) cohort, a project funded by the National Institutes of Health (NIH) and launched in 2004 (adni.loni.usc.edu)^25^. Individuals had available information on cortical and subcortical brain regions volume, cerebrospinal fluid (CSF) biomarkers and genetic data. Participants were classified into A/T groups defined by their CSF biomarker profile according to the A/T framework described in^26^. Amyloid-beta pathology positivity (A+) and tau pathology positivity (T+) were defined based on validated cut-offs^27^. Further details on the CSF sampling can be found in <u>Supplementary Methods</u>. The CSF profile of A+T+ individuals in ALFA was determined by the CSF Aβ42/40 ratio < 0.071 pg/mL and levels of CSF p-tau181 > 0.24 pg/mL, while in ADNI A+ was defined by CSF Aβ42 levels ≤1100 pg/mL. Participants were classified into four different groups based on both their clinical diagnosis and their CSF amyloid profile. Finally, the sample was covering the whole disease *continuum*: CU A-ALFA (N=220), CU A+ ALFA (N=118), MCI A+ ADNI (N=230), AD A+ ADNI (N=100).

### 2.2. Genetic data acquisition, quality control and imputation

For the ALFA study, DNA was obtained from blood samples through a salting-out protocol. Genotyping was performed with the Illumina Infinium Neuro Consortium (NeuroChip) Array (build GRCh37/hg19)^28^. Quality control procedure was performed using PLINK software. Imputation was performed using the Michigan Imputation Server with the haplotype Reference Consortium Panel (HRC r1.1 2016;^29^ following default parameters and established guidelines. A full description of the genotyping, quality control and imputation procedures is available elsewhere^13^. ADNI participants underwent genotyping using both the Human 610-Quad BeadChip and the Illumina HumanOmniExpress BeadChip (Illumina, Inc., San Diego, CA). Quality control measures and imputation procedures closely followed the protocols established in the ALFA study. Further details are available elsewhere^30^.

### 2.3. Genetic predisposition to Alzheimer’s disease: polygenic risk score computation

The polygenic risk score of AD (PRS_AD_) was calculated using the PRSice version 2 tool^31^. Summary statistics from a recent GWAS for AD^11^ were obtained to compute the PRS ^13^ [Supplementary Table 1]. The algorithm retained the single nucleotide polymorphisms (SNPs) with the smallest p-value in each 250 kb window and removed SNPs that were in linkage disequilibrium (r2>0.1). The threshold of SNPs inclusion was defined at p-value<5·10^-6^. The PRS_AD_ was computed by adding up the alleles carried by participants, weighted by the SNP allele effect size from the GWAS and normalizing by the total number of alleles. The same procedure was performed to estimate the PRS of AD when excluding the *APOE* region (PRS_ADnoAPOE_) (chr19:45,409,011-45,412,650; GRCh37/hg19). PRSs were both computed in ALFA and ADNI, respectively^13^. Both PRSs were dichotomized and we created two groups of subjects to differentiate individuals at higher genetic predisposition to AD from the rest of the sample. Categorization was done in each cohort separately. The cut-off point was defined by the quantile 0.8 (High genetic group: PRS≥quantile 0.8; Low genetic group: PRS<quantile 0.8).

### 2.4. Brain regions segmentation and quantification

Volumes for cortical and subcortical brain regions were obtained through high-resolution 3D-T1-weighted MRI scans. Each cohort used customized protocols specific to the scanner. The images were pre-processed and segmented using Freesurfer. In ALFA^32^, protocol parameters were identical for all participants and included high-resolution three-dimensional structural images weighted in T1 with an isotropic voxel of 0.75 × 075 × 0.75 mm^3^, acquired with a 3T Philips ingenia CS scanner The acquisition parameters were TR/TE/TI = 9.9/4.6/900 ms, flip angle = 8° and a matrix size of 320 × 320 × 240. In ADNI, MRI acquisition parameters included a repetition time (TR) of 2,400 ms, inversion time (TI) of 1,000 ms, flip angle of 8 degrees, and a field of view (FOV) of 24 cm. The acquisition matrix was set to 256 × 256 × 170 in the x-, y-, and z-dimensions, resulting in a voxel size of 1.25 × 1.26 × 1.2 mm ^33^. The MRI data underwent preprocessing steps, including correction for anterior commissure and posterior commissure, skull stripping, cerebellum removal, intensity inhomogeneity correction. Finally, the data were down-sampled to a matrix size of 128 × 128 × 128. Freesurfer version 7 was used for cortical and subcortical quantification in ALFA, while in ADNI, version 5 and version 6 were used in different batches. Cortical surface parcellation was done using the Desikan-Killiany atlas^34^. Subcortical measurements were obtained working with the automatic subcortical segmentation of Freesurfer. Volumes were globally quantified by summing the measurements of both hemispheres. A total of 41 volumes were selected combining 34 measurements from cortical regions and 7 from subcortical ones [<u>Supplementary Methods</u>]. Harmonization across sites was not assessed because the compositional approach did not work with raw volumes of the individuals. Instead, it worked with all pairs of log-ratios between regions’ volumes. Therefore, any differences that could arise between cohorts were removed.

### 2.5. Statistical analyses

Differences in demographic characteristics were assessed according to amyloid status (ALFA) and diagnosis group (ADNI), using chi-square tests for categorical variables and parametric (t-test, ANOVA) and non-parametric tests (Wilcoxon Rank-Sum test or Kruskal-Wallis test) for continuous normally and non-normally distributed variables, respectively. P-values for pairwise comparisons were also provided, adjusted for Benjamini-Hochberg false discovery rate. CoDA ^35–22^ was performed to identify structural brain signatures (i.e. a combination of specific brain regions’ volumes) (i) capable of distinguishing between CU A- and other groups along the disease *continuum*, and (ii) associated with higher genetic predisposition to AD within groups [<u>Supplementary Methods</u>]. In the first scenario, logistic regression models, adjusted for age, sex and the PRS_AD_, were defined to examine the structural brain signature that distinguished CU A-from the rest of the groups in the *continuum* (CU A+, MCI A+, AD A+). In the second step, disease-stage stratified logistic regression models, adjusted for age and sex, were performed to explore the association between the brain signature with higher genetic risk of AD in each group [Figure 1]. Separate analyses were performed based on the PRS calculation used to determine the genetic groups: 1) The PRS including all the SNPs associated with AD (PRS_AD_) and 2) The PRS excluding the *APOE* region (PRS_ADnoAPOE_). The procedure was exactly the same and analyses were repeated for both cases. Results for the second case can be found in <u>Supplementary Results</u>. All the analyses were performed using the R software version 4.2.2.

**Figure 1.**
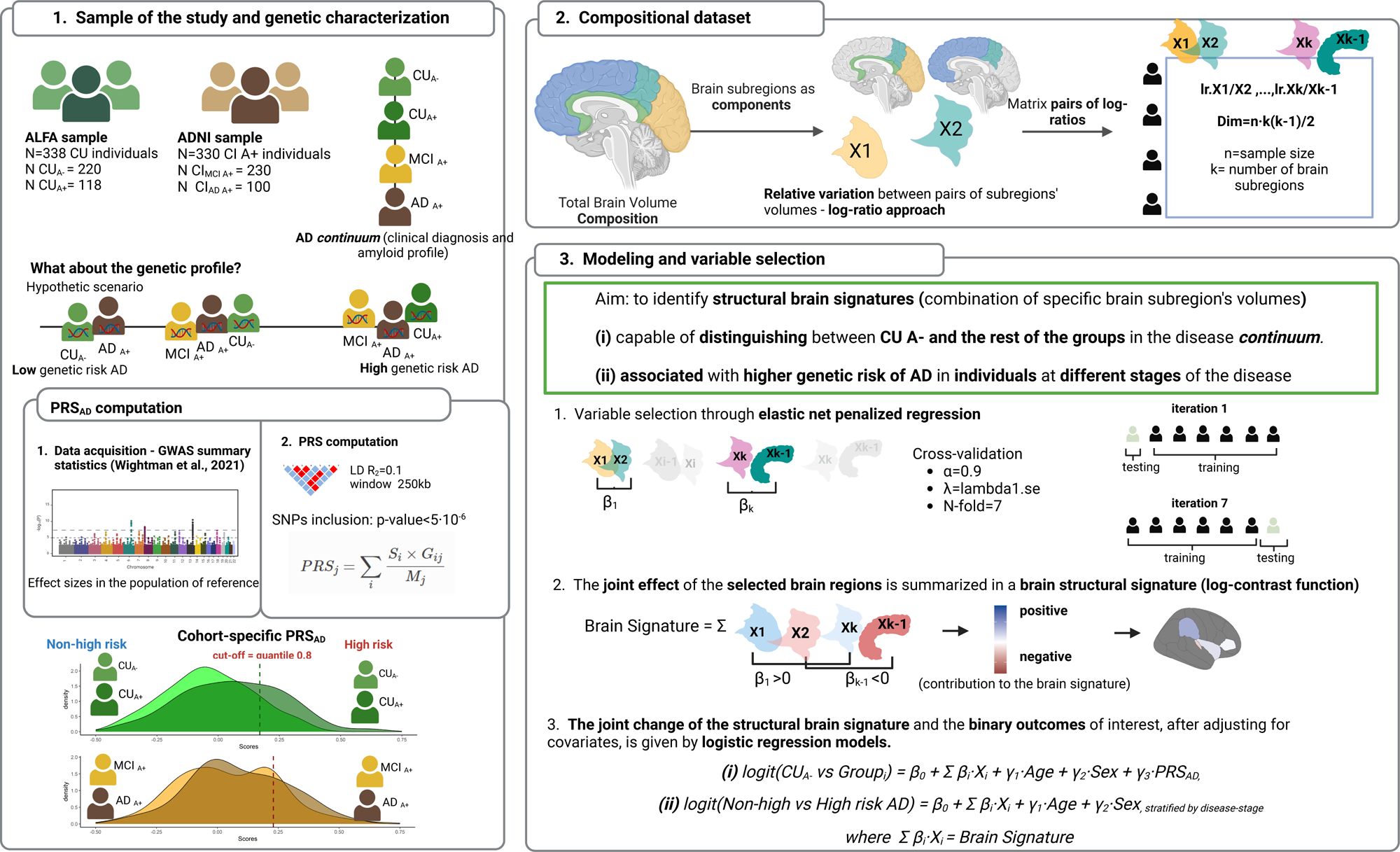
Workflow *coda4microbiome* algorithm: implementation in our Imaging Genetics study.

## 3. Results

### 3.1. Characteristics of the sample

The ALFA sample (N=338) included 62.43% of women and 54.14% of *APOE-ε4* carriers, with a median age of 57 years old [IQR 53-61][Table 1]. The ADNI sample (N=330) was defined by 40% of women and ∼64.55% of *APOE-ε4* carriers, with a median age of 74 years [IQR 68.2-78]. In the sample there were 30% of AD dementia patients and 70% of MCI individuals, all of them amyloid positive [Table 2]. Differences were assessed among disease-stage groups along the *continuum* [Table 3]. The highest percentage of women was found in ALFA CU A-individuals (63%) while the lowest was observed in ADNI MCI individuals (38%) (p < 0.001). Significant differences in *APOE-ε4* carriership were observed among groups, with the lowest percentage found in ALFA CU A-participants (40% carriers) and the highest in ALFA CU A+ (78% carriers) (p < 0.001). Notably, the *APOE-ε4* carrier rate in ALFA CU A+ was higher than in clinical groups from ADNI (p < 0.001). There were significant differences in the genetic predisposition to AD between CN A- and CU A+, MCI and AD dementia individuals (FDR p-value pairwise comparisons < 0.001), but not between CU A+ and the impaired groups. Non-significant differences were found between groups when the *APOE* region was removed from the polygenic risk score [Supplementary Figure 1]. Significant differences were also found in the median value of cortical and subcortical regions between groups [Supplementary Table 2].

**Table 1.** Demographic characteristics and APOE genotypes distribution in the ALFA sample.

|  | **ALFA**<br>**N=338** | **N** |
| --- | --- | --- |
| **Sex (n,%)** |  | 338 |
| **Men** | 127 (37.574%) |  |
| **Women** | 211 (62.426%) |  |
| **Age (median, IQR)** | 57.000 [53.000;61.000] | 338 |
| **Education years (median, IQR)** | 12.000 [11.000;17.000] | 338 |
| **Amyloid-beta status (n,%)** |  | 338 |
| **AB-** | 220 (65.089%) |  |
| **AB+** | 118 (34.911%) |  |
| **APOE ε4 carriers (n,%)** |  | 338 |
| **Non-carrier** | 155 (45.858%) |  |
| **Carrier** | 183 (54.142%) |  |
| **APOE ε4 load (n,%)** |  | 338 |
| **Non-carriers** | 155 (45.858%) |  |
| **One ε4 allele** | 152 (44.970%) |  |
| **Two ε4 alleles** | 31 (9.172%) |  |
| **Minimental State Examination (median, IQR)** | 29.000 [28.000;30.000] | 338 |

**Table 2.** Demographic characteristics and APOE genotypes distribution in the ADNI sample.

|  | **ADNI**<br>**N=330** | **N** |
| --- | --- | --- |
| **Sex (n,%)** |  | 330 |
| **Women** | 132 (40.000%) |  |
| **Men** | 198 (60.000%) |  |
| **Age (median, IQR)** | 74.050 [68.200;78.000] | 330 |
| **Education years (median, IQR)** | 16.000 [14.000;18.000] | 330 |
| **Diagnosis (n,%)** |  | 330 |
| **AD** | 100 (30.303%) |  |
| **MCI** | 230 (69.697%) |  |
| **APOE ε4 carriers (n,%)** |  | 330 |
| **Non-carrier** | 117 (35.455%) |  |
| **Carrier** | 213 (64.545%) |  |
| **APOE ε4 load (n,%)** |  | 330 |
| **Non-carriers** | 117 (35.455%) |  |
| **One ε4 allele** | 153 (46.364%) |  |
| **Two ε4 alleles** | 60 (18.182%) |  |
| **Minimental State Examination (median, IQR)** | 27.000 [25.000;29.000] | 330 |

**Table 3.**
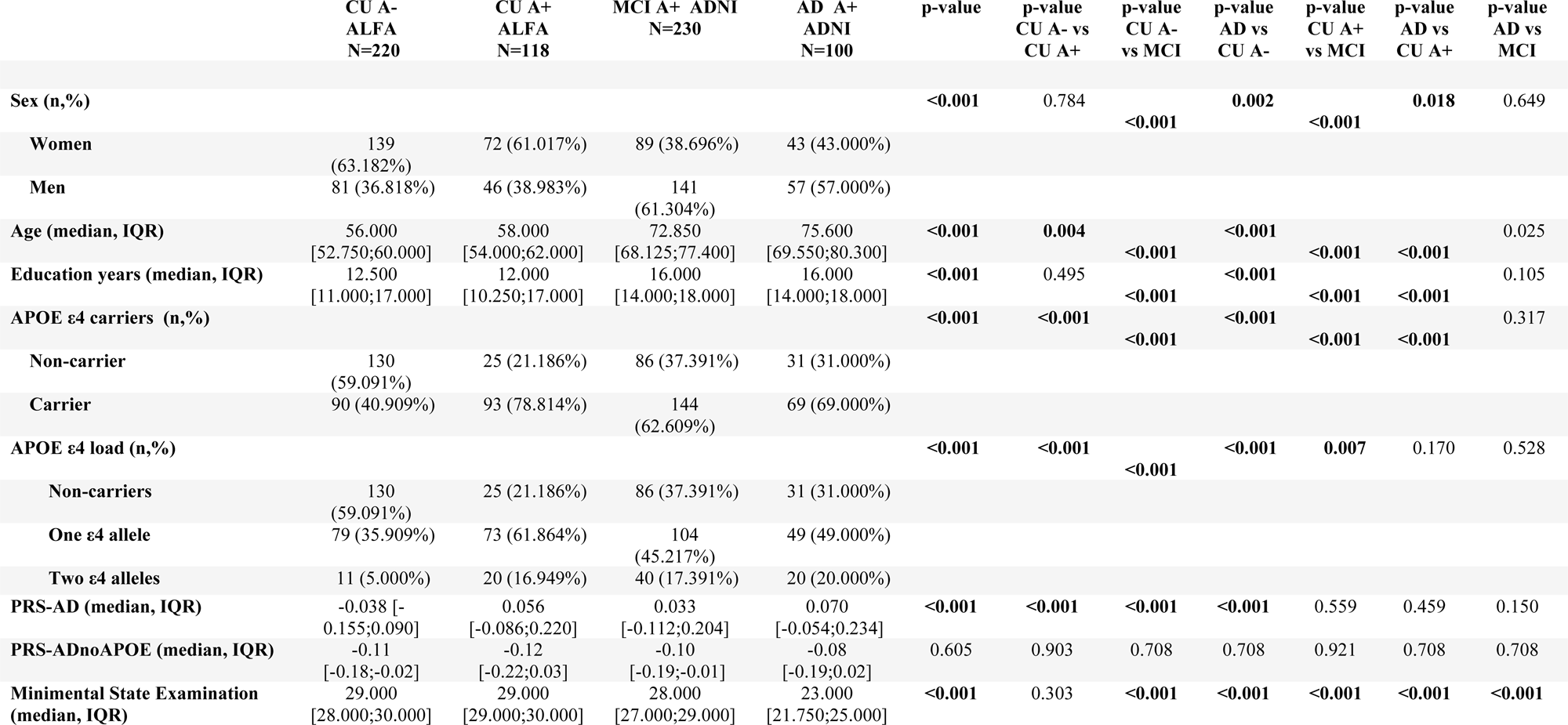
Demographic characteristics and APOE genotypes distribution in the ALFA and ADNI sample.

### 3.2. Structural brain signatures differentiate amyloid-negative cognitively unimpaired individuals from others at different stages of the disease

In the ALFA study, the structural brain signature that differentiated CU A-from CU A+ individuals was primarily characterized by the relative volumetric variation between the pallidum and the amygdala, along with volumetric variations in other cortical regions of the parietal, frontal and temporal lobes. The brain signature also involved subcortical regions, specifically the pallidum, putamen and the amygdala, and well as cortical areas in the cingulate cortex [Figure 2]. Thee structural brain signature capable of distinguishing between CU A- and MCI A+ individuals mainly involved the relative volumetric variation between the fusiform and the frontal pole, along with volumetric variations of other temporal and frontal areas, as well as regions in the occipital lobe. This signature also included subcortical regions, specifically the putamen and pallidum, and cortical areas in the cingulate [Figure 2]. Finally, the structural brain signature that distinguished between CU A- and AD A+ individuals was primarily characterized by the relative volumetric variation between the insula and the inferior parietal, along with the volumetric variation of other parietal and frontal regions. The brain signature also involved the putamen and cortical areas in the cingulate cortex [Figure 2]. The structural brain signature showed high prediction accuracy for differentiating CU A-from disease stages, with the discrimination being significantly robust for both MCI A+ (AUC=0.992) and AD A+ (AUC=0.980) [Supplementary Table 3].

**Figure 2.**
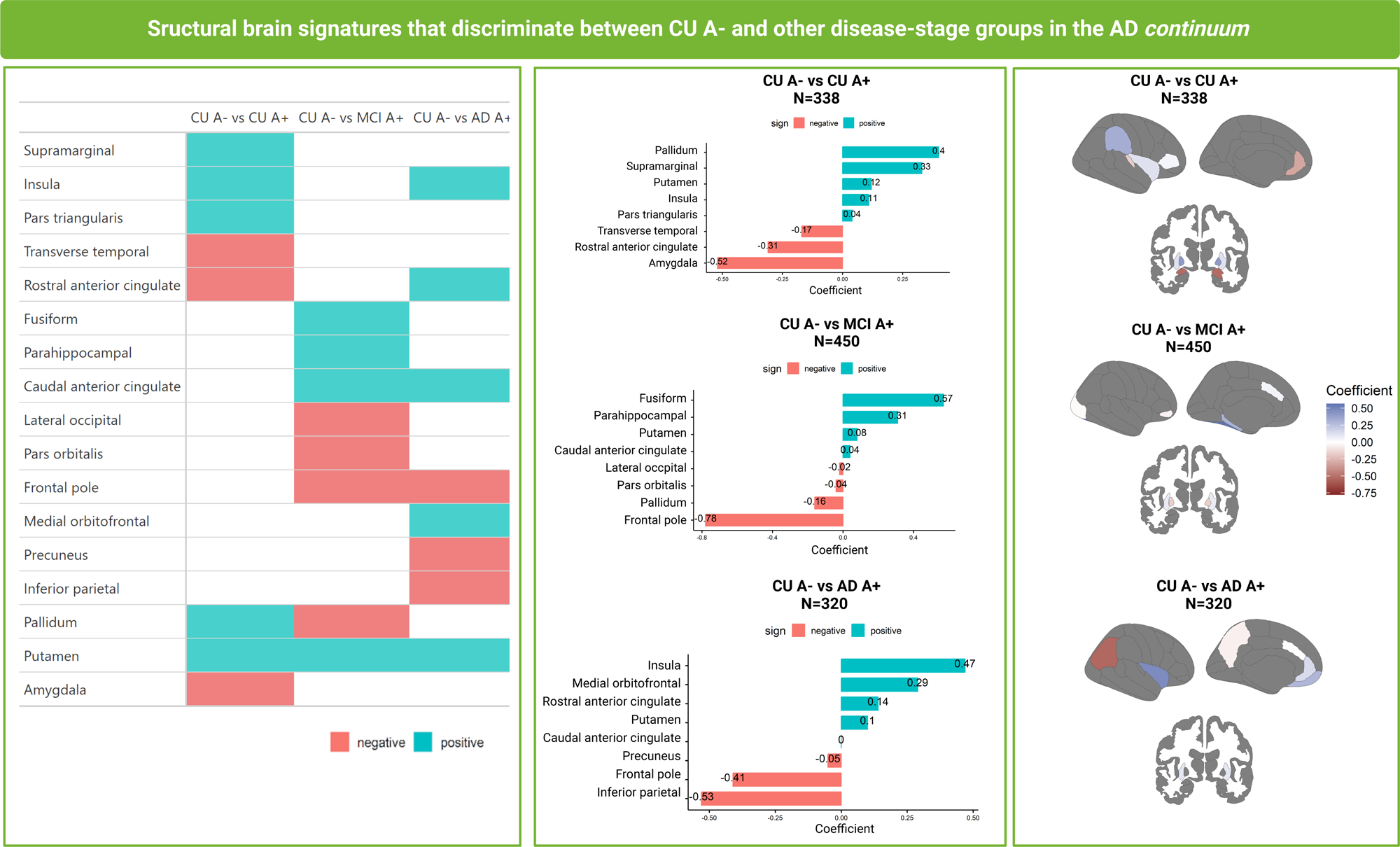
Brain structural signature distinguishing between CU A-individuals and others at different stages of the disease. In blue, brain regions that positively contribute to the structural brain signature. In red, brain regions that negatively contribute to the structural brain signature. *Legend: CU A-(cognitively unimpaired amyloid-beta negative individuals), CU A+ (cognitively unimpaired amyloid-beta positive individuals), MCI A+ (mild cognitive impaired amyloid-beta positive individuals), AD A+ (Alzheimer’s disease amyloid-beta positive individuals)*.

### 3.3. Disease-stage specific structural brain signatures associated with higher genetic risk of Alzheimer’s disease

In CU A-individuals, the structural brain signature associated with higher genetic risk of AD primarily featured structural changes in the hippocampus and the fusiform, along with variations in frontal, temporal and occipital areas together with subcortical regions such as the pallidum [Figure 3]. For CU A+ individuals, the structural brain signature related to higher risk of AD was characterized by volumetric variations in temporal, frontal and occipital regions, with a higher emphasis on the temporal areas, specifically the middle and inferior temporal regions [Figure 3]. When exploring these structural brain signatures in CI individuals, we observed that in MCI participants, the structural brain signature was primarily defined by structural changes in the pars opercularis and the superior parietal, together with variations in other frontal and parietal regions, along with contributions from some temporal and occipital areas as well as subcortical regions such as the amygdala and area accumbens [Figure 3]. In AD dementia individuals, the structural brain signature associated with higher genetic risk of AD was mainly characterized by structural changes in the pars triangularis and the amygdala, along with variations in other frontal regions, as well as parietal, temporal and occipital areas [Figure 3D]. Although the prediction accuracy of the structural brain signature was low in all the groups [Supplementary Table 4], there were significant differences in the mean score of the structural brain signature between at-high and at-low risk individuals in each group along the disease *continuum* [Supplementary Figure 3].

**Figure 3.**
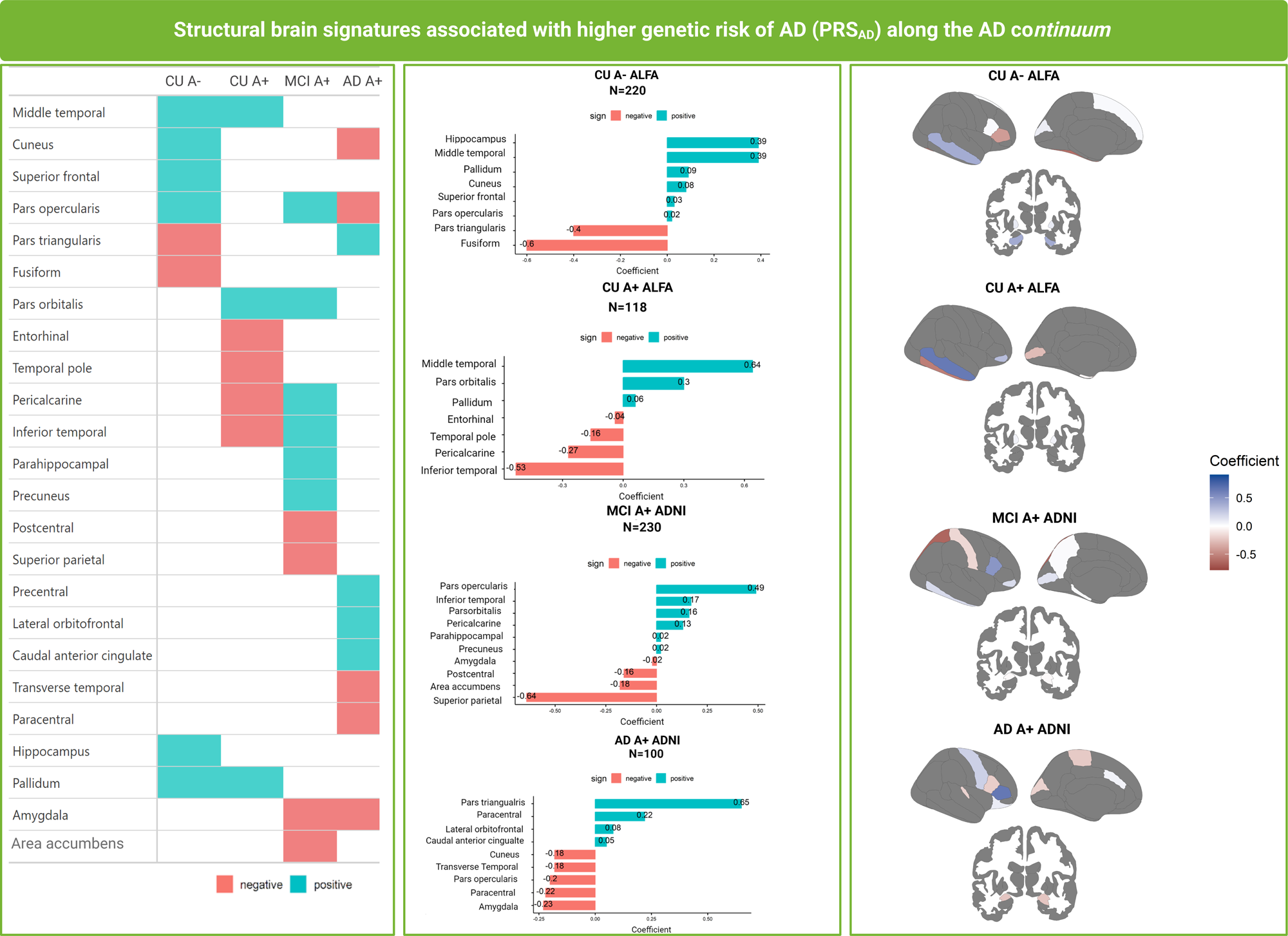
Brain structural signature associated with higher genetic predisposition to AD (PRS_AD_). In blue, brain regions that positively contribute to the structural brain signature. In red, brain regions that negatively contribute to the structural brain signature. *Legend: CU A-(cognitively unimpaired amyloid-beta negative individuals), CU A+ (cognitively unimpaired amyloid-beta positive individuals), MCI A+ (mild cognitive impaired amyloid-beta positive individuals), AD A+ (Alzheimer’s disease amyloid-beta positive individuals)*.

## 4. Discussion

In this study, the use of CoDA revealed structural brain signatures capable of discerning between CU A-individuals and subjects within other disease-stage groups along the AD *continuum*. We also found structural brain signatures associated with higher genetic predisposition to AD that were specific to each stage of the disease along the *continuum*. In both cases, the structural brain signatures reflected different structural trajectories among the groups.

On the one hand, regions involved in the brain signatures discriminating between CU A- and the rest of the groups were in line with previous reported patterns of brain networks in CU, MCI and AD^36^. The structural brain signature discerning between CU A- and CU A+ primarily involved subcortical areas such as the amygdala and pallidum, and cortical areas in the insula, cingulate and both frontal and parietal lobes. Despite being inversely related, the amygdala and pallidum emerged as the regions with the most significant contribution to the structural brain signature. Some authors ^37^ have found that atrophy in the amygdala predicts Mini Mental State Examination scores and hippocampal atrophy in very mild and mild AD subjects. These findings highlight the relationship between the magnitude of atrophy in this region and cognitive impairment in very early stages of the disease. The structural brain signature that distinguished between CU A- and MCI A+, was mainly characterized by cortical regions in the temporal, frontal and occipital lobe, as well as in the cingulate cortex, together with subcortical areas such as the putamen and pallidum. Despite their inverse relationship, the fusiform and the frontal pole showed the most pronounced contribution to the structural brain signature. Recent studies have revealed different patterns of atrophy in MCI individuals, with observed thinning in both the fusiform and the frontal in distinct incident MCI subtypes^38^. Results highlight the complexity of structural changes in heterogeneous profiles. Finally, CU A-individuals were differentiated from AD A+ through a structural brain signature mainly involving cortical areas in the frontal and parietal lobe, together with the insula and the cingulate cortex, as well as subcortical areas such as the putamen. The insula and the inferior parietal were inversely related but had the highest contribution to the structural brain signature. Previous studies have explored cortical networks in CU, MCI and AD patients^36^ and have identified group-specific hub regions, specifically involving the cingulate and the orbital frontal gyrus in AD patients. These findings are in line with the reported regions that we found involved in the structural brain signature associated with higher odds of belonging to the AD group. Moreover, authors ^36^ also found that compared with CU, the nodal centrality of MCI and AD decreased in the middle temporal and increased in the precuneus, located in the parietal lobe.

On the other hand, regions involved in the structural brain signatures associated with higher genetic predisposition of AD within disease-stage groups, reflected the structural atrophy trajectory observed as AD progresses, from medial temporal, to temporo-parietal and frontal regions^39^. The structural brain signature linked to higher risk of AD in CU individuals primarily involved the relative change of temporal areas. Furthermore, independently of amyloid status, the structural brain signature of both CU A- and A+ individuals, shared some commonalities, such as the involvement of the pallidum and the middle temporal cortex, albeit with varying degrees of prominence. Despite these common similarities, the impact of AD genetic load on the modulation of nearby brain regions was group-specific. These findings suggest that the brain’s response to AD genetic predisposition differs even within the preclinical stage, which has been already observed in univariate brain modelization^17–18^. In the MCI group, the structural brain signature discerning individuals at higher genetic risk of AD from those at lower risk, was characterized by relative volumetric changes in temporal and parietal regions. Specifically, MCI at higher genetic risk of AD exhibited a brain signature representing an intermediate position between CU and AD. Notably, the amygdala and accumbens played significant roles in shaping this signature. The amygdala, in particular, is well-established in the processing of emotional and memory-related information, implying a potential role in the early stages of cognitive dysfunction (Al-Ani et al., 2023). In the AD group, the structural brain signature was primarily characterized by frontal regions. The amygdala remained a crucial contributor to this signature. These findings emphasize the genetic susceptibility of the amygdala and its role in advanced stages of AD. Recent studies^40^ have reported the association between the *APOE-ε4* genotype and reduced volumes in the amygdala, specifically in the AD dementia and MCI groups compared to young cognitively healthy individuals. However, they did not find significant associations between the volume of the amygdala and an overall polygenic score of AD. In contrast, other studies^41^ have found significant associations between higher genetic risk of AD and lower amygdala volumes in individuals from the ADNI cohort.

Altogether, results suggested that when accounting for genetic information, group-specific structural brain signatures better capture the pattern of structural changes observed as the disease progresses. The identification of these structural brain signatures underscores the complex interplay between genetics and brain morphology in both prodromal and clinical stages of the disease. Moreover, these findings provide a novel perspective for brain IG studies showing that the contribution of genetic factors to brain structure may evolve as the disease progresses^42^. Results also highlighted the impact of polygenicity on the brain morphology. While the *APOE*-*ε4* allele is widely recognized as the most significant risk factor for AD, our study exposes the importance of a broader genetic landscape in influencing brain structure alterations in AD^43^. This insight is particularly crucial for the understanding of the disease pathogenesis, suggesting that changes in brain structure are a result of a complex genetic interplay rather than being solely dependent on single genetic variants^44^. Besides the *APOE-ε4* allele, other polymorphisms in the *APOE* region (*i.e.* rs405509; T/T as risk allele), have been independently associated with the thickness of specific brain regions (e.g. left parahippocampal gyrus) in old cognitively healthy individuals^45^. Moreover, genetic variants included in the computation of the polygenic risk score of AD were annotated to multiple genes ^11^ that have been associated with MRI outcomes, such as *TOMM40*^46^, that is involved in a haplotype together with *APOE* (TOMM40′523-*APOE* ε4) that has been associated with AD cortical morphological traits ^47^.

Despite these findings, our study also showed some limitations. Notably, we acknowledge the need for longitudinal data analysis to assess the joint volumetric trajectories discerning individuals at higher genetic risk of AD from those at lower risk over time. Another aspect to consider is that each structural brain signature captures the relative variation of brain regions’ volumes that are most closely associated with the outcome of interest. Thus, the structural brain signature does not reflect actual changes on the brain volumetric composition, but relative volumetric variations. This interpretation might not be straightforward, but the methodology overcomes biases and limitations of current IG multivariate approaches when working with heterogeneous MRI data from different cohorts. For instance, CoDA surpasses traditional methodologies in its ability to integrate volumetric data from various cohorts without the stringent requirement for harmonization^48^. This flexibility facilitates the analysis of diverse datasets and strengthens the reliability of cross-study comparisons. Nonetheless, another limitation lies in the involvement of other AD risk factors that may interact with genetics affecting the brain volumetric modulation. It is well known that genetic factors significantly contribute to neurodegenerative disorders, but often they are functional under specific exposures^49^. In this context, one could state that individuals from the ALFA cohort are exposed to sufficiently homogeneous conditions to assume an equal random impact of factors external to genetics. However, it should be noted that an extension of these results by evaluating the impact of external factors and in a more diverse population group, would improve the interpretation and translation of the results.

This study also had several strengths. The use of CoDA represents a strong method for overcoming methodological aspects of the traditional analyses used in neuroimaging studies and offers a more accurate and comprehensive understanding of brain structural alterations linked to genetics than common univariate and multivariate methods^50^. Unlike conventional univariate analyses, which often lack the power to detect significant associations due to rigorous multiple testing corrections at brain-wide or genome-wide level, this approach adeptly captures the complex interdependencies among different brain regions. Moreover, CoDA offers distinct advantages over commonly used multivariate methods^19^. While multivariate approaches reduce data dimensionality and identify correlated feature sets^51^, they typically overlook the compositional nature of brain data.

In summary, CoDA emerges as a relevant method in neuroimaging studies, holding significant implications for both research and clinical practice. By providing a more accurate analysis of brain imaging data and addressing its compositional nature, CoDA can influence the development of targeted approaches, opening new avenues for enhancing brain health.

## Data Availability

All data produced in the present study are available upon reasonable request to the authors.

## Acknowledgements

This publication is part of the ALFA study. The authors express their most sincere gratitude to the ALFA project participants and relatives without whom this research would have not been possible. Collaborators of the ALFA study are: Müge Akinci, Federica Anastasi, Annabella Beteta, Raffaele Cacciaglia, Lidia Canals, Alba Cañas, Carme Deulofeu, Maria Emilio, Irene Cumplido-Mayoral, Marta del Campo, Carme Deulofeu, Ruth Dominguez, Maria Emilio, Karine Fauria, Sherezade Fuentes, Marina García, Laura Hernández, Gema Huesa, Jordi Huguet, Laura Iglesias, Esther Jiménez, David López-Martos, Paula Marne, Tania Menchón, Paula Ortiz-Romero, Eleni Palpatzis, Wiesje Pelkmans, Albina Polo, Sandra Pradas, Mahnaz Shekari, Lluís Solsona, Anna Soteras, Núria Tort-Colet, and Marc Vilanova.

Data collection and sharing for this project was funded by the Alzheimer’s Disease Neuroimaging Initiative (ADNI) (National Institutes of Health Grant U01 AG024904) and DOD ADNI (Department of Defense award number W81XWH-12-2-0012). ADNI is funded by the National Institute on Aging, the National Institute of Biomedical Imaging and Bioengineering, and through generous contributions from the following: AbbVie, Alzheimer’s Association; Alzheimer’s Drug Discovery Foundation; Araclon Biotech; BioClinica, Inc.; Biogen; Bristol-Myers Squibb Company; CereSpir, Inc.; Cogstate; Eisai Inc.; Elan Pharmaceuticals, Inc.; Eli Lilly and Company; EuroImmun; F. Hoffmann-La Roche Ltd and its affiliated company Genentech, Inc.; Fujirebio; GE Healthcare; IXICO Ltd.; Janssen Alzheimer Immunotherapy Research & Development, LLC.; Johnson & Johnson Pharmaceutical Research & Development LLC.; Lumosity; Lundbeck; Merck & Co., Inc.; Meso Scale Diagnostics, LLC.; NeuroRx Research; Neurotrack Technologies; Novartis Pharmaceuticals Corporation; Pfizer Inc.; Piramal Imaging; Servier; Takeda Pharmaceutical Company; and Transition Therapeutics. The Canadian Institutes of Health Research is providing funds to support ADNI clinical sites in Canada. Private sector contributions are facilitated by the Foundation for the National Institutes of Health (www.fnih.org). The grantee organization is the Northern California Institute for Research and Education, and the study is coordinated by the Alzheimer’s Therapeutic Research Institute at the University of Southern California. ADNI data are disseminated by the Laboratory for Neuro Imaging at the University of Southern California.

## Consent Statement

The ALFA study was conducted in accordance with the directives of the Spanish Law 14/2007, of July 3, on Biomedical Research (Ley 14/2007 de Investigación Biomédica). The ALFA study protocol was approved by the Independent Ethics Committee Parc de Salut Mar Barcelona and registered at Clinicaltrials.gov (Identifier: NCT01835717). All participants accepted the study procedures by signing the study’s informed consent form that had also been approved by the same Institutional Review Board.

