## Supplementary material for "Compositional structural brain signatures capture Alzheimer’s genetic risk on brain structure along the disease *continuum*": SF1.pdf

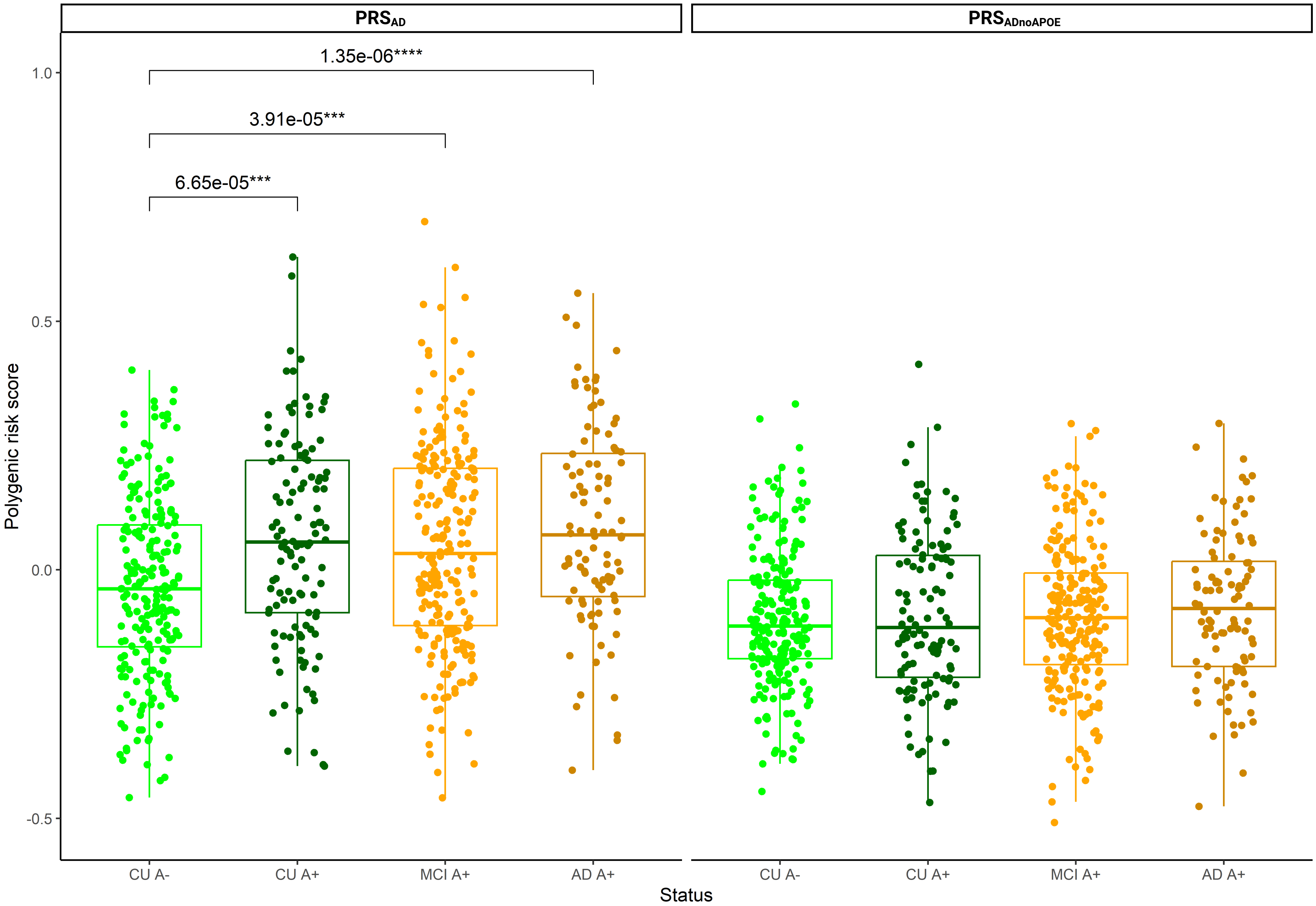


**Supplementary Figure 1.** Distribution of genetic scores of Alzheimer's disease along the AD continuum. Pairwise comparisons are assessed to compare the median $PRS_{AD}$ (Wilcoxon test) among groups. *Significant results at FDR p-value are displayed (FDR p-value <0.05 *; FDR p-value <0.01 **, FDR p-value <0.001 ***).*
