## Supplementary material for "Compositional structural brain signatures capture Alzheimer’s genetic risk on brain structure along the disease *continuum*": SF4.pdf

### Structural brain signatures associated with higher genetic risk of AD when excluding the *APOE* region ($PRS_{ADnoAPOE}$) along the AD *continuum*

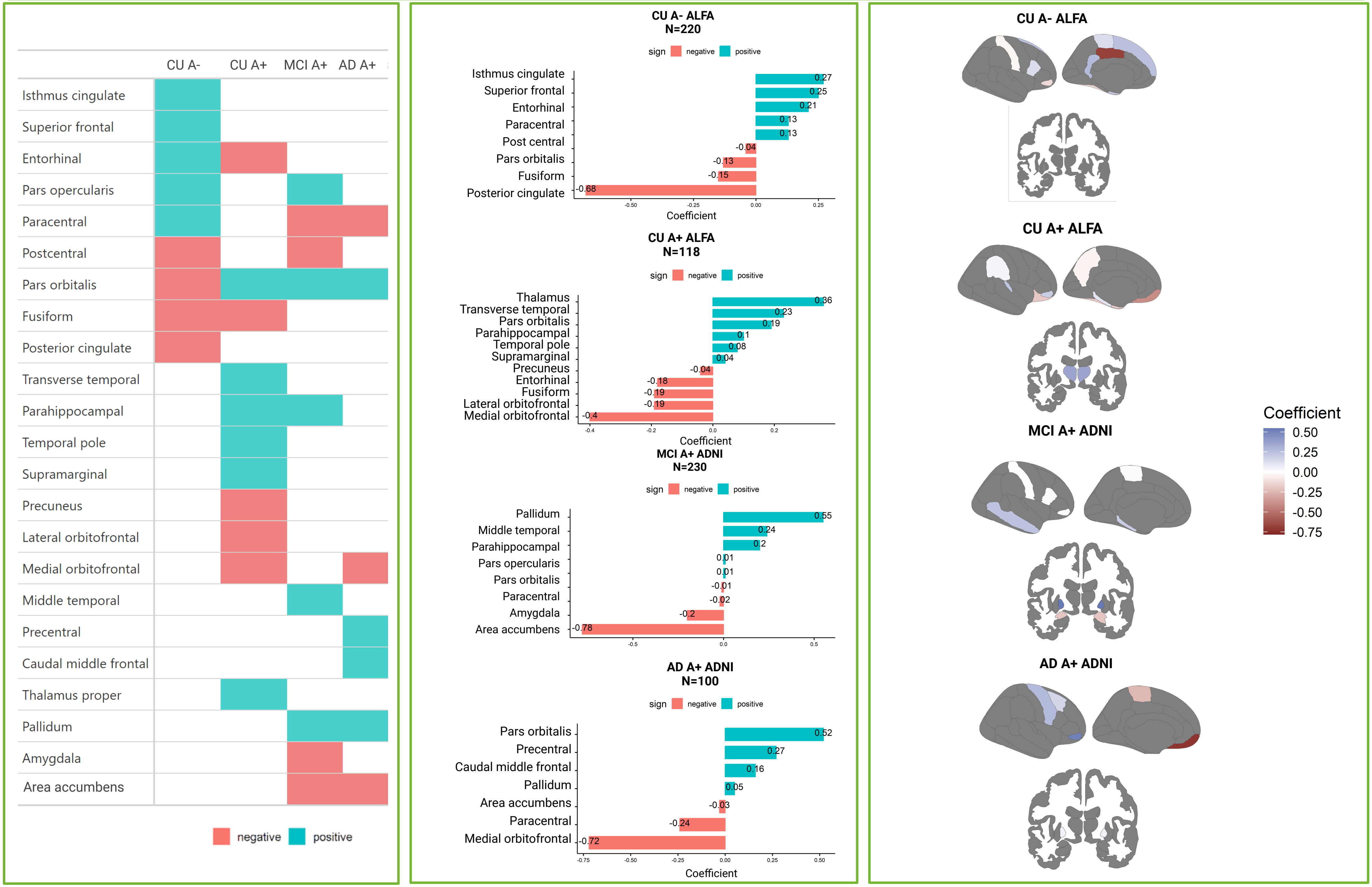


**Supplementary Figure 4.** Brain regions' contribution to the structural brain signature associated with higher genetic predisposition to AD after excluding the *APOE* region ($PRS_{ADnoAPOE}$). In blue, brain regions that positively contribute to the structural brain signature. In red, brain regions that negatively contribute to the structural brain signature. *Legend: CU A- (cognitively unimpaired amyloid-beta negative individuals), CU A+ (cognitively unimpaired amyloid-beta positive individuals), MCI A+ (mild cognitive impaired amyloid-beta positive individuals), AD A+ (Alzheimer's disease amyloid-beta positive individuals).*
