## Supplementary material for "Compositional structural brain signatures capture Alzheimer’s genetic risk on brain structure along the disease *continuum*": SF5.pdf

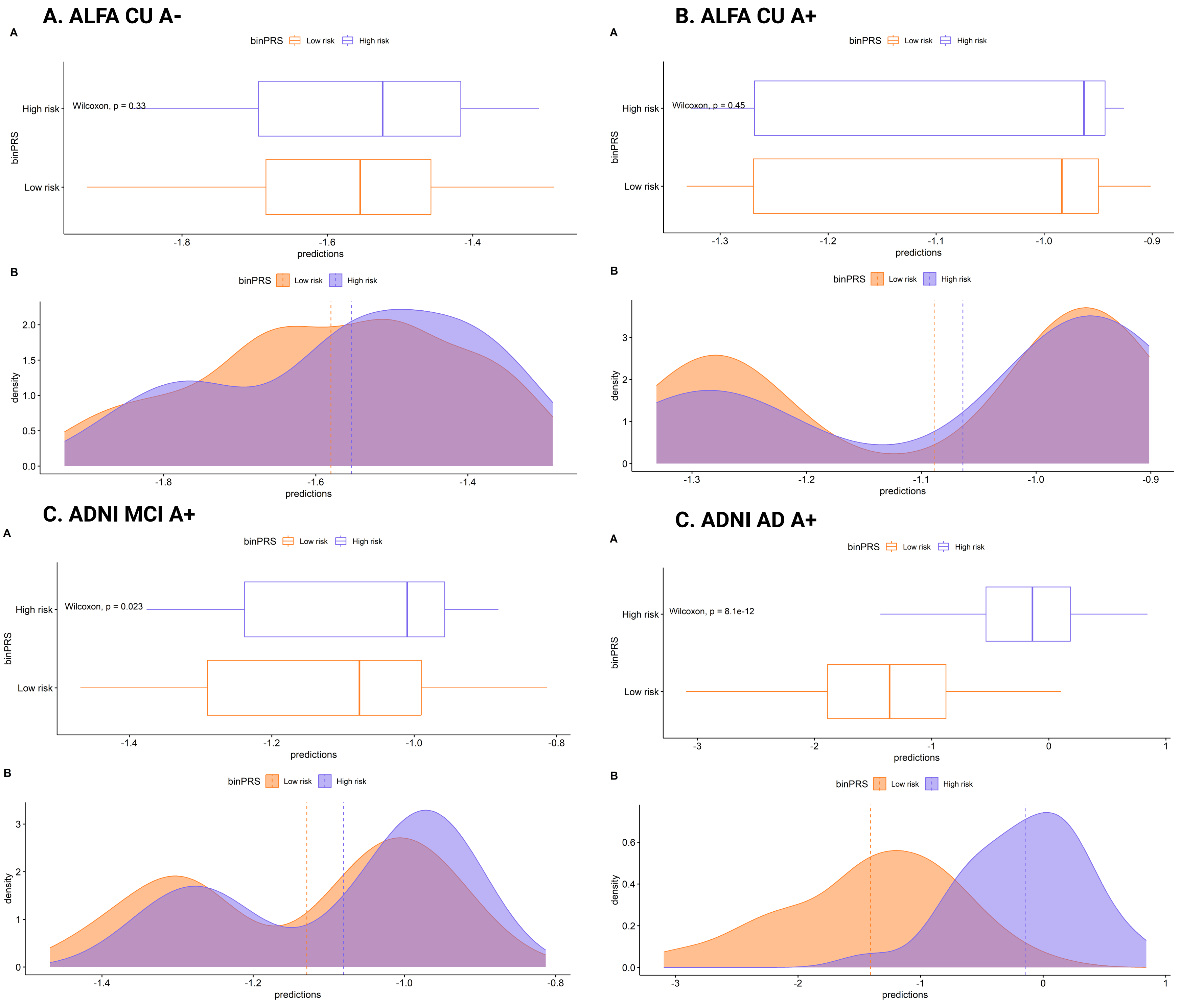


**Supplementary Figure 5.** Prediction plot describing the distribution of the predicted scores of the structural brain signature within disease-stage groups, stratifying by AD genetic risk. Median values of the predicted scores were compared between groups (p-values for Wilcoxon Rank-Sum test).
