## Supplementary material for "Compositional structural brain signatures capture Alzheimer’s genetic risk on brain structure along the disease *continuum*": Supplementary Methods.docx

*Cerebrospinal fluid data sampling and analysis*

A subset of the ALFA study participants were invited to take part in a nested longitudinal long-term study, referred to as ALFA+, in which more detailed phenotyping is performed. It entails the acquisition of both fluid (CSF, blood) and imaging (MRI and PET) biomarkers, as well as an extended cognitive assessment. Specifically, 419 individuals have been included and undergone ALFA+ baseline visit (October 2016 - December 2019), with the first follow-up visit currently ongoing (November 2019 - (expected) Q2 2022). Of them, 393 with available CSF and genetic data, and 286 with available PET and genetic data are included in the ALFA study. CSF samples were obtained by lumbar puncture following standard procedures (Milà-Alomà et al., 2020; Teunissen et al., 2014), and all the measurements were conducted at the Clinical Neurochemistry Laboratory, Sahlgrenska University Hospital, Mölndal, Sweden. Aβ pathology positivity (A+) and tau pathology positivity (T+) were defined by CSF Aβ42/40 ratio and CSF Mid(M)p-tau181, respectively (Jack et al. 2016). The derived cut-offs are explained in detail in (Milà-Alomà et al. 2020) and were used to define AT groups (i.e. A+T+. A+T, A-T-).

*Compositional data analysis applied to Brain Imaging Genetics*

Compositional data consists of a vector of positive measurements whose values are restricted by their total sum (Calle 2019) (**Equation 1**). The value of each component is not informative by itself and the relevant information is contained in the ratios between the parts.

**(Equation 1**) $x=[x_{1},....,x_{D}] \in R^{D},$

for $x_{1}>0, \sum_{i=1}^{D} x_{i}=k$, where *k* is a constant

The present study aimed to explore (i) the capability of 41 cortical and subcortical brain segmentations’ volumes of compositional origin to distinguish between groups at different stages of the *continuum* (**Equation 2)** as well as (ii) the association between brain segmentation’s volumes of compositional origin and higher genetic risk of AD (**Equation 3)**, adjusting for covariates in both cases.

(**Equation 2)** $logit(CU_{A-} vs Group_{i}) = \beta_{0} + \sum_{i=1}^{D} \beta_{i} x_{i} + \gamma_{1}Age +\gamma_{2}Sex +\gamma_{3}PRS_{AD}$,

where *i* $\in(CU_{A+}, MCI_{A+}, AD_{A+})$

(**Equation 3)** $logit(Non-high vs High risk AD) = \beta_{0} + \sum_{i=1}^{D} \beta_{i} x_{i} + \gamma_{1}Age +\gamma_{2}Sex$

Commonly, compositional methods are based on a log-ratio approach to assure scale invariance. In this work we implemented a new methodology, known as *coda4microbiome* (Calle et al., 2023), that involves both modeling and variable selection. This approach looks for the variables (components) most closely associated with the response variable of interest. In the modeling step, either a logistic or a linear regression model is considered depending on the outcome variable of interest (**Equation 3**).

**(Equation 3**) $logit(Y) = \beta_{0} + \beta_{1}x + \gamma Z$, when Y is binary

$Y= \beta_{0} + \beta_{1}x + \gamma Z$, when Y is continuous

All pairs of log-ratios between components are included in the model as predictors (**Equation 4)** (Calle et al., 2023).

(**Equation 4)** $g(E(y))=\beta_{0}+\sum_{1\leq j<k\leq K} \beta_{jk}\cdot log(x_{j}/x_{k})$

Variable selection is obtained through elastic net penalized regression and the penalty term (lambda; 𝛌) is defined through a n-fold cross-validation procedure.

The signature is defined as the linear combination of the selected log-ratios **(Equation 5**) .

(**Equation 5**) Signature = $\sum_{1\leq j<k\leq K} \hat{\beta}_{jk}\cdot log(x_{j}/x_{k})$

After reparameterization, the signature can be expressed as a weighted sum of the selected variables in the form of a log-contrast function (Calle et al., 2023). The signature can be interpreted as a balance between two groups of variables: those with a positive contribution and those with a negative one (Susin et al., 2020) (**Equation 6**).

**(Equation 6**) $\sum_{j=1}^{K} \hat{\theta}_{j}\cdot log(x_{j})$, where $\hat{\theta}_{j}=\sum_{k=j+1}^{K} \hat{\beta}_{jk} -\sum_{j=1}^{K} \hat{\beta}_{kj}$ and $\sum_{j=0}^{K} \hat{\theta}_{j}=0$

*Description of the brain regions*

| **Component** | **Anatomical Region** | **Lobe** | **Region** |
| --- | --- | --- | --- |
| v1 | Cortical | Frontal | Superior Frontal |
| v2 | Cortical | Frontal | Rostral |
| v3 | Cortical | Frontal | Caudal Middle Frontal |
| v4 | Cortical | Frontal | Pars Opercularis |
| v5 | Cortical | Frontal | Pars Orbitalis |
| v6 | Cortical | Frontal | Pars Triangularis |
| v7 | Cortical | Frontal | Lateral orbitofrontal |
| v8 | Cortical | Frontal | Medial Orbitofrontal |
| v9 | Cortical | Frontal | Precentral |
| v10 | Cortical | Frontal | Paracentral |
| v11 | Cortical | Frontal | Frontal Pole |
| v12 | Cortical | Parietal | Superior Parietal |
| v13 | Cortical | Parietal | Inferior Parietal |
| v14 | Cortical | Parietal | Supramarginal |
| v15 | Cortical | Parietal | Postcentral |
| v16 | Cortical | Parietal | Precuneus |
| v17 | Cortical | Temporal | Superior Temporal |
| v18 | Cortical | Temporal | Middle Temporal |
| v19 | Cortical | Temporal | Inferior Temporal |
| v20 | Cortical | Temporal | Banks |
| v21 | Cortical | Temporal | Fusiform |
| v22 | Cortical | Temporal | Transverse Temporal |
| v23 | Cortical | Temporal | Entorhinal |
| v24 | Cortical | Temporal | Temporal Pole |
| v25 | Cortical | Temporal | Parahippocampal |
| v26 | Cortical | Occipital | Lateral Occipital |
| v27 | Cortical | Occipital | Lingual |
| v28 | Cortical | Occipital | Cuneus |
| v29 | Cortical | Occipital | Pericalcarine |
| v30 | Cortical | Cingulate | Rostral anterior |
| v31 | Cortical | Cingulate | Caudal anterior |
| v32 | Cortical | Cingulate | Posterior |
| v33 | Cortical | Cingulate | Isthmus |
| v34 | Cortical | Insular Cortex | Insula |
| v35 | Subcortical | Temporal | Amygdala |
| v36 | Subcortical | Temporal | Hippocampus |
| v37 | Subcortical | Basal Ganglia | Pallidum |
| v38 | Subcortical | Basal Ganglia | Putamen |
| v39 | Subcortical | Diencephalon | Thalamus |
| v40 | Subcortical | Basal Ganglia | Caudate |
| v41 | Subcortical | Ventral Striatum | Area accumbens |
