## Supplementary material for "Compositional structural brain signatures capture Alzheimer’s genetic risk on brain structure along the disease *continuum*": Supplementary Results.docx

*Structural brain signatures associated with higher genetic risk of AD, excluding the APOE region, in the disease continuum*

When the *APOE* region was removed from the genetic score influencing the classification of at-risk CU A- individuals, the brain signature associated with higher genetic risk of AD was mainly characterized by structural changes in the cingulate cortex along with variations in frontal regions [[Supplementary Figure 4A](https://drive.google.com/file/d/13mRs1DmZaWlR4pcBGe-O9JIPGZQ1xk4P/view?usp=sharing)]. In CU A+ individuals, the brain signature associated with higher risk was defined by subcortical structural changes in thalamus together with volumetric variations in temporal (e.g. transverse temporal, parahippocampal region, temporal pole) and frontal regions (e.g. lateral and medial orbitofrontal) [[Supplementary Figure 4B](https://drive.google.com/file/d/13mRs1DmZaWlR4pcBGe-O9JIPGZQ1xk4P/view?usp=sharing)].

In MCI A+ individuals, the brain signature was mainly defined by subcortical structural changes in the pallidum, amygdala and area accumbens along with the variation of temporal regions [[Supplementary Figure 4C](https://drive.google.com/file/d/13mRs1DmZaWlR4pcBGe-O9JIPGZQ1xk4P/view?usp=sharing)]. In AD A+ individuals, the brain signature associated with higher genetic risk of AD was mainly characterized by cortical structural changes in frontal regions together with subcortical changes in the pallidum [[Supplementary Figure 4D](https://drive.google.com/file/d/13mRs1DmZaWlR4pcBGe-O9JIPGZQ1xk4P/view?usp=sharing)]. Although the prediction accuracy of the brain signature was low in all the groups [[Supplementary Table 5](https://docs.google.com/spreadsheets/d/11LjxBReK1O1jE3RN-w9h_WbpcWmZH4OTq7w4B8LtUaM/edit" \l "gid=0)], there were significant differences in the mean score of the brain signature between at-high risk individuals and the rest of the subjects in the group of MCI A+ and AD A+ [[Supplementary Figure 5C-D](https://drive.google.com/file/d/1EPg35WgMBTqup8oF0X8TTiAB2-zW_0hw/view?usp=sharing)].
